## Supplementary Material 1 for "*APOE4* and Infectious Diseases Jointly Contribute to Brain Glucose Hypometabolism, a Biomarker of Alzheimer’s Pathology: New Findings from the ADNI"

**Supplementary Materials**

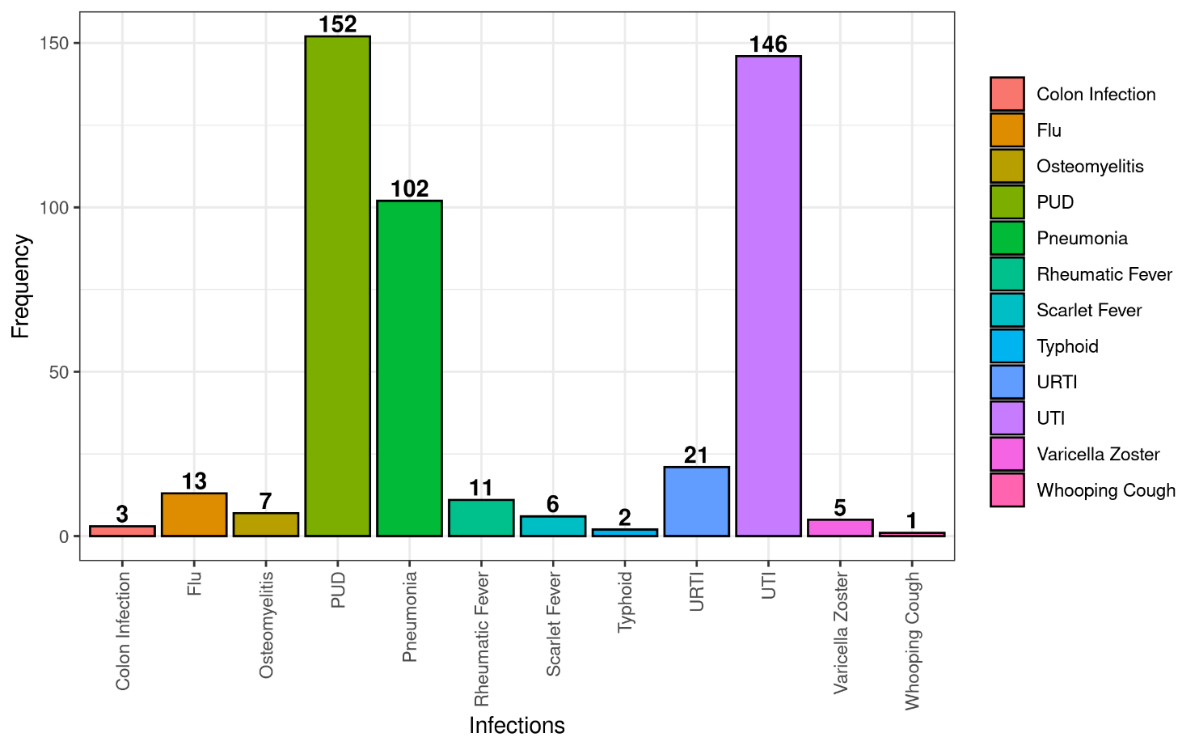

Supplementary Fig 1. Types of infections and their frequencies (n = 218, infections = 242)

Note. The same individual had multiple infections on a few occasions. *Abbreviations:* PUD, Peptic Ulcer Disease; URTI, Upper Respiratory Tract Infection; UTI, Urinary Tract Infection.

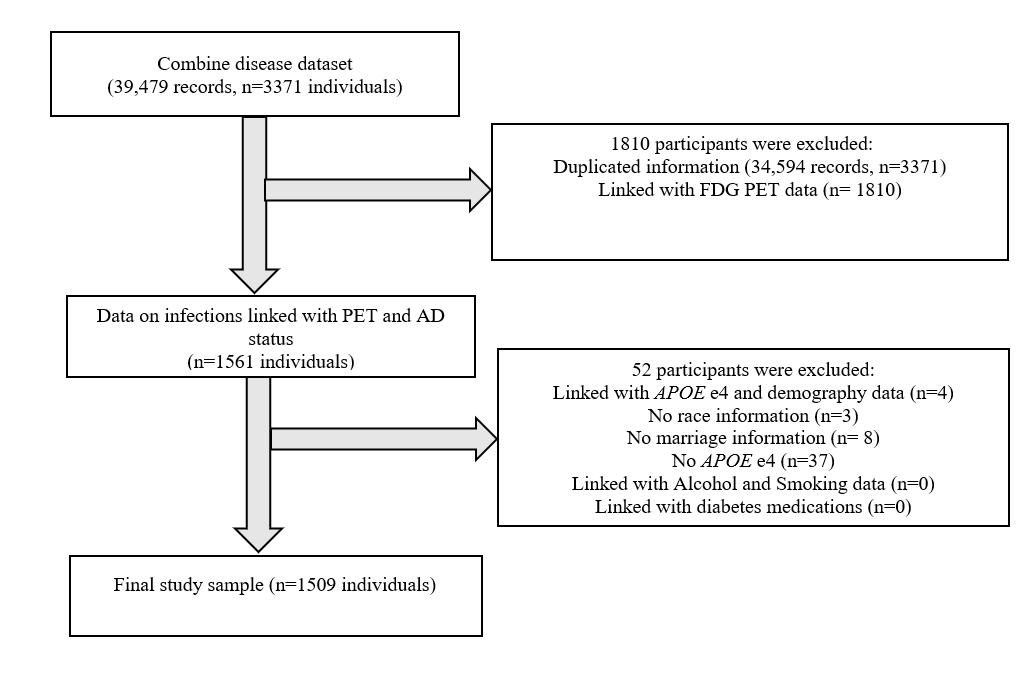

Supplementary Fig 2. Study flowchart of participant selection

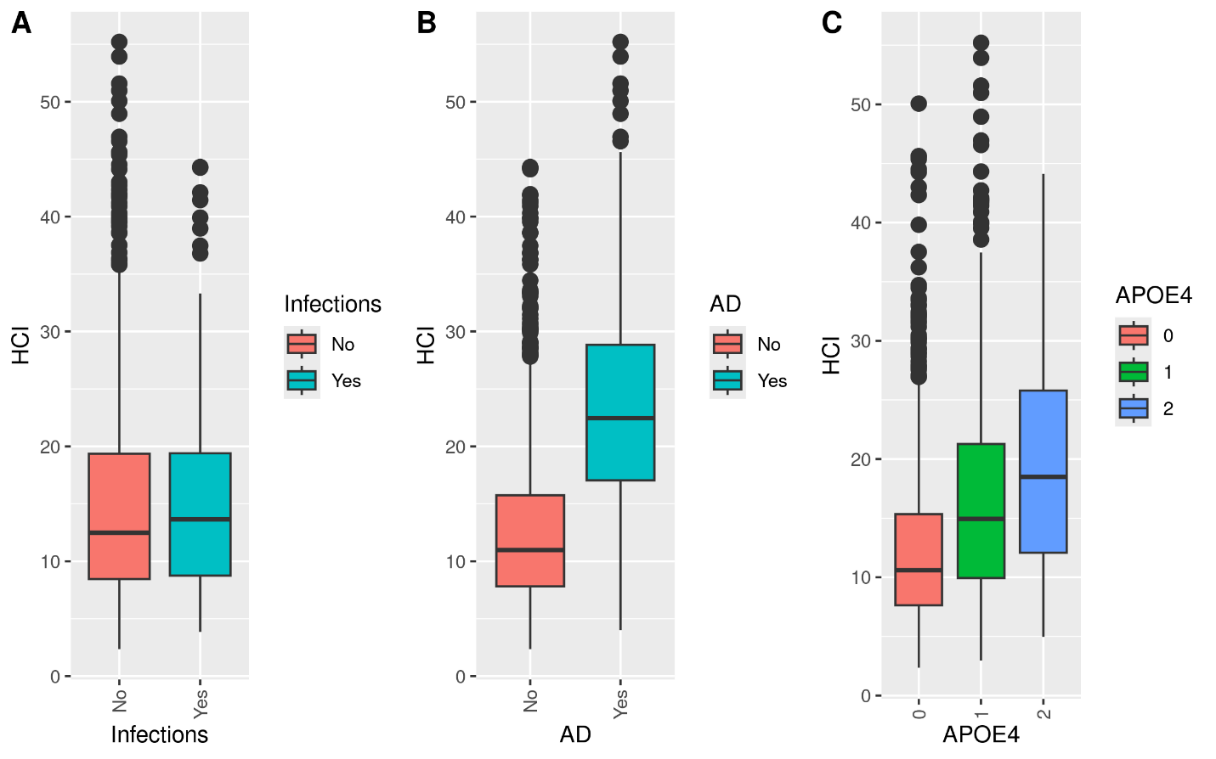

Supplementary Fig 3. Distribution of HCI values (A) Distribution for individuals with infections (B) Distribution for individuals with Alzheimer's disease (AD) (C) Distribution for individuals with *APOE4*

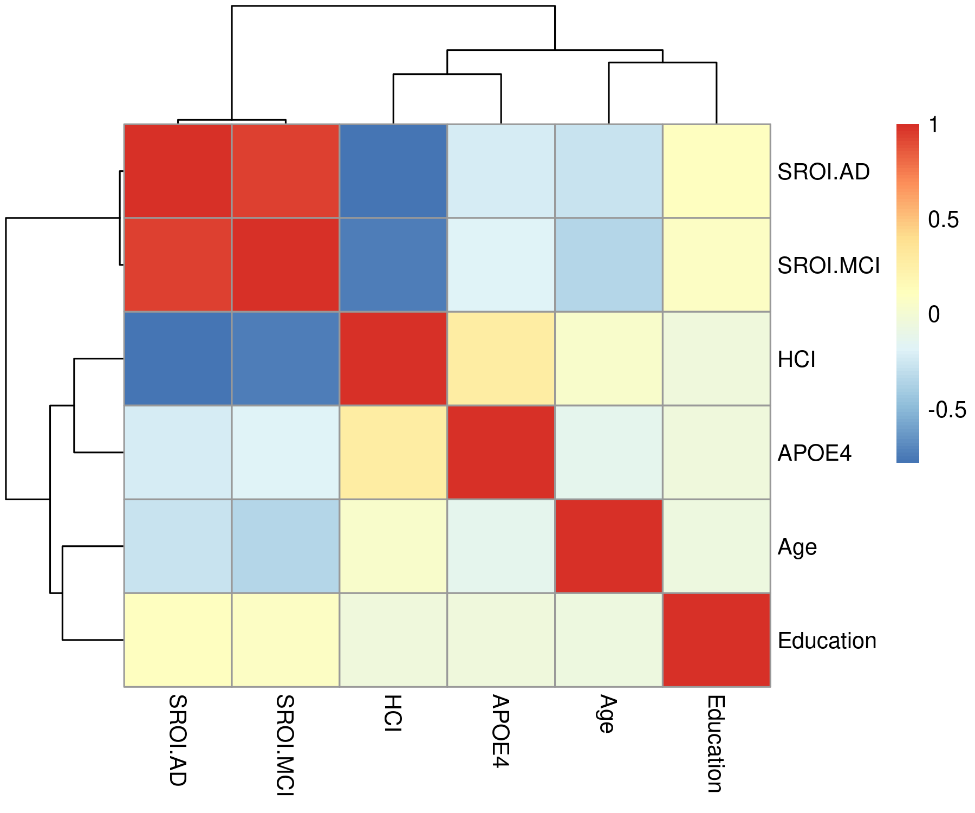

Supplementary Fig 4. Correlation between continuous covariates with investigated brain metabolism measures.

Note. The darker shades indicate strong positive or negative correlations.

Supplementary Table 1. Regression estimates for predictors in the multivariate linear regression full model for HCI outcome

| **Variables** | **Estimates** | **95% CI** | **p** |
| --- | --- | --- | --- |
| Infections (Yes) | 0.15 | 0.03 -0.28 | 0.01^*^ |
| Age | 0.01 | 0.01, 0.02 | <0.001^***^ |
| Sex (Male) | 0.17 | 0.08, 0.26 | <0.001^***^ |
| Education (Years) | -0.002 | -0.02, 0.01 | 0.81 |
| Marriage (Never) | -0.06 | -0.29, 0.17 | 0.61 |
| *APOE4* | 0.32 | 0.25, 0.38 | <0.001^***^ |
| Race (White) | 0.25 | 0.09, 0.42 | 0.002^**^ |
| Smoking (Yes) | 0.08 | -0.01, 0.19 | 0.095 |
| Alcohol (Yes) | -0.02 | -0.27, 0.23 | 0.873 |
| AD (Yes) | 1.04 | 0.92, 1.16 | <0.001^***^ |
| Diabetes Medication (Yes) | 0.19 | -0.07, 0.45 | 0.159 |

Note. ^*^p<0.05; ^**^p<0.01; ^***^p<0.001.

Supplementary Table 2. Measures of variable importance computed using Random forest models

| **Variable** | **%IncMSE** | **IncNodepurity** |
| --- | --- | --- |
| AD | 81.59 | 22222.41 |
| *APOE4* | 36.10 | 6778.73 |
| Age | 13.72 | 10642.90 |
| Sex | 12.31 | 1466.75 |
| Race | 9.67 | 1021.57 |
| Smoking | 4.54 | 1048.22 |
| Infections | 0.63 | 1014.08 |

Note. %IncMSE represents the increase in the regression mean-squared error when variables are randomly permuted. IncNodePurity measures the loss function based on the optimal node splits that maximize variance. Higher values for both metrics indicate more important variables.

Supplementary Table 3. Marginal means showing the model-predicted HCI stratified by sex

| **Infections** | **Sex** | **Mean age** | **Age range** | **Marginal**  **means** | **SE** | **95% CI** | **p-value** |
| --- | --- | --- | --- | --- | --- | --- | --- |
| No | Female | 72.23 | 55.0 - 89.6 | 0.26 | 0.05 | 0.15, 0.37 | <0.001^***^ |
| Yes | Female | 72.92 | 60.0 - 90.3 | 0.41 | 0.07 | 0.25, 0.56 | <0.001^***^ |
| No | Male | 73.95 | 55.0 - 91.4 | 0.43 | 0.05 | 0.33, 0.54 | <0.001^***^ |
| Yes | Male | 75.65 | 57.8 - 89.3 | 0.58 | 0.07 | 0.44, 0.73 | <0.001^***^ |

Note. The means are averaged over the levels of age, sex, race, smoking, and AD.

Supplementary Table 4. Marginal means showing the model-predicted HCI stratified by *APOE4*

| **Infections** | ***APOE4*** | **Marginal means** | **SE** | **95% CI** | **p-value** |
| --- | --- | --- | --- | --- | --- |
| No | 0 | 0.03 | 0.05 | -0.07, 0.13 | 0.53 |
| Yes | 0 | 0.18 | 0.07 | 0.03, 0.33 | 0.01^*^ |
| No | 1 | 0.38 | 0.05 | 0.27, 0.49 | <0.001^***^ |
| Yes | 1 | 0.53 | 0.07 | 0.38, 0.68 | <0.001^***^ |
| No | 2 | 0.62 | 0.08 | 0.47, 0.78 | <0.001^***^ |
| Yes | 2 | 0.77 | 0.09 | 0.58, 0.97 | <0.001^***^ |

Note. The means are averaged over the levels of age, sex, race, smoking, and AD.

Supplementary Table 5. Association of number of infections with HCI

| **Variables** | **Estimates** | **95% CI** | **p** |
| --- | --- | --- | --- |
| Infections (n=1) | 0.11 | -0.01, 0.24 | 0.08 |
| Infections (n>=2) | 0.44 | 0.09, 0.80 | 0.01^*^ |
| Age | 0.01 | 0.006, 0.01 | <0.001^***^ |
| Sex (Male) | 0.17 | 0.08, 0.25 | <0.001^***^ |
| *APOE4* | 0.31 | 0.25, 0.38 | <0.001^***^ |
| Race (White) | 0.25 | 0.09, 0.41 | 0.002^**^ |
| Smoking (Yes) | 0.08 | -0.01, 0.18 | 0.09 |
| AD (Yes) | 1.03 | 0.92, 1.15 | <0.001^***^ |

Note. ^*^p<0.05; ^**^p<0.01; ^***^p<0.001.

Supplementary Table 6. Regression estimates for predictors in the multivariate linear regression full model for SROI AD outcome

| **Variables** | **Estimates** | **95% CI** | **p** |
| --- | --- | --- | --- |
| Infections (Yes) | -0.01 | -0.02, -0.001 | 0.02^*^ |
| Age | -0.003 | -0.003, -0.002 | <0.001^***^ |
| Sex (Male) | -0.01 | -0.01, -0.002 | 0.009^**^ |
| Education (Years) | 0.001 | 0.000, 0.002 | 0.043^*^ |
| Marriage (Never) | 0.01 | -0.01, 0.03 | 0.26 |
| APOE4 | -0.02 | -0.03, -0.02 | <0.001^***^ |
| Race (White) | -0.01 | -0.02, 0.004 | 0.18 |
| Smoking (Yes) | -0.01 | -0.02, 0.001 | 0.10 |
| Alcohol (Yes) | 0.01 | -0.01, 0.03 | 0.52 |
| AD (Yes) | -0.08 | -0.09, -0.07 | <0.001^***^ |
| Diabetes Medication (Yes) | -0.03 | -0.05, -0.005 | 0.01^**^ |

Note. ^*^p<0.05; ^**^p<0.01; ^***^p<0.001.

Supplementary Table 7. Regression estimates for predictors in the multivariate linear regression full model for SROI MCI outcome

| **Variables** | **Estimates** | **95% CI** | **p** |
| --- | --- | --- | --- |
| Infections (Yes) | -0.013 | -0.026, -0.0005 | 0.04^*^ |
| Age | -0.004 | -0.005, -0.004 | <0.001^***^ |
| Sex (Male) | -0.022 | -0.032, -0.013 | <0.001^***^ |
| Education (Years) | 0.001 | -0.0005, 0.002 | 0.19 |
| Marriage (Never) | 0.007 | -0.016, 0.031 | 0.52 |
| *APOE4* | -0.022 | -0.029, -0.016 | <0.001^***^ |
| Race (White) | -0.005 | -0.022, 0.010 | 0.49 |
| Smoking (Yes) | -0.006 | -0.016, 0.0038 | 0.22 |
| Alcohol (Yes) | -0.003 | -0.028, 0.022 | 0.81 |
| AD (Yes) | -0.090 | -0.102, -0.078 | <0.001^***^ |
| Diabetes Medication (Yes) | -0.035 | -0.061, -0.008 | <0.01^**^ |

Note. ^*^p<0.05; ^**^p<0.01; ^***^p<0.001.
